# Participant perceptions and experiences of a novel community-based pilot respiratory longitudinal sampling method in Liverpool, UK

**DOI:** 10.1101/2023.05.18.23289716

**Authors:** E.L. German, H.M. Nabwera, R. Robinson, F. Shiham, K. Liatsikos, C.M. Parry, C. McNamara, S. Kattera, K. Carter, A. Howard, S. Pojar, J. Hamilton, A. Matope, J. Read, S.J. Allen, H. Hill, D.B. Hawcutt, B.C. Urban, A.M. Collins, D.M. Ferreira, E. Nikolaou

**Affiliations:** Department of Clinical Sciences, Liverpool School of Tropical Medicine, Liverpool, UK; Alder Hey Children’s Hospital, Liverpool, UK; Centre of Excellence for Women and Child Health, Aga Khan University, Nairobi, Kenya; Liverpool University Hospitals NHS Foundation Trust, Liverpool, UK; Lancaster Medical School, Lancaster University, Lancaster, UK; Edward Francis Small Teaching Hospital, Banjul, The Gambia; University of Liverpool, Liverpool, UK; Oxford Vaccine Group, Department of Paediatrics, University of Oxford, Oxford, UK; Infection and Immunity, Murdoch Children’s Research Institute, Melbourne, Victoria, Australia; Department of Microbiology and Immunology, Peter Doherty Institute for Infection and Immunity, University of Melbourne, Melbourne, Victoria, Australia

**Author notes:** **Corresponding author:** Dr Elissavet Nikolaou, Murdoch Children’s Research Institute, Parkville, Victoria, Australia, 3050. The last three authors contributed equally to the manuscript.

## Abstract

Longitudinal, community-based sampling is important for understanding prevalence and transmission of respiratory pathogens. Using a minimally invasive sampling method, the FAMILY Micro study monitored the oral, nasal and hand microbiota of families for 6 months. Here, we explore participant experiences and opinions.

A mixed methods approach was utilised. A quantitative questionnaire was completed after every sampling timepoint to report levels of discomfort and pain, as well as time taken to collect samples. Participants were also invited to discuss their experiences in a qualitative structured exit interview.

We received questionnaires from 36 families. Most adults and children >5y experienced no pain (94% and 70%) and little discomfort (73% and 47% no discomfort) regardless of sample type, whereas children ≤5y experienced variable levels of pain and discomfort (48% no pain but 14% hurts even more, whole lot or worst; 38% no discomfort but 33% moderate, severe, or extreme discomfort). The time taken for saliva and hand sampling decreased over the study.

We conducted interviews with 24 families. Families found the sampling method straightforward, and adults and children >5y preferred nasal sampling using a synthetic absorptive matrix over nasopharyngeal swabs. It remained challenging for families to fit sampling into their busy schedules. Adequate fridge/freezer space and regular sample pick-ups were found to be important factors for feasibility. Messaging apps proved extremely effective for engaging with participants.

Our findings provide key information to inform the design of future studies, specifically that self-sampling at home using minimally invasive procedures is feasible in a family context.

## Introduction

Respiratory tract infections are a leading cause of morbidity and mortality among post-neonatal children under five years worldwide [1, 2]. Although microorganisms in the respiratory tract can cause serious disease, especially in the young, old, and immunocompromised, they are also often carried asymptomatically [3-5], creating reservoirs for transmission throughout the population. Age, coinfections, and underlying health conditions are known to affect rates and density of asymptomatic carriage and infection [5-9].

Community-based longitudinal studies are hugely beneficial for understanding the epidemiology of respiratory tract infections and the host-pathogen interactions involved in pathways to disease. However, such studies are impeded by sampling methods like nasopharyngeal swabs, which are invasive and unpleasant, particularly for children [10-12]. In addition, families are often required to travel to health care/research facilities or to schedule home visits for sample collection.

Previous studies have shown that self-collected and/or parent-collected swabs are feasible and effective methods for studying respiratory tract infections such as influenza and RSV [13-16]. Comparing healthcare professional (HCP) taken swabbing to self-swabbing, the latter had higher participation rates, lower costs and showed equal sensitivity in identification of bacteria favouring this method for use in large population-based respiratory carriage studies [17]. Self-testing at home has become more popular with the recent COVID-19 pandemic, as it alleviates work pressures and infection risk for healthcare workers [18]. There is also increased interest in using saliva for respiratory sampling, both for bacteria [19, 20] and viruses [21, 22] as it is much easier to collect with minimal discomfort to the patient/participant.

Our group has successfully established a home sampling methodology, collecting saliva and nasal lining fluid for studying pneumococcal carriage after experimental exposure in adults [23]. The Family Research of Microbes Linked to Respiratory Infections (FAMILY Micro) study expands our approach to community-based research and assesses the acceptability and feasibility of our methodology for surveillance of respiratory pathogens among parents and their children. Our approach was acceptable and well-received by the participants [24]. Here, we explored participant experiences of the FAMILY Micro study, assessing the feasibility of our methodology for future application in large-scale surveillance studies.

## Methods

### Overview of the FAMILY Micro study

Full details of the study recruitment, conduct and its primary outcomes (acceptability and compliance) are described elsewhere [24]. Briefly, the FAMILY Micro study (ISCTRN 52814289, NHSREC 284708) ran between October 2020 and August 2021 and was a mixed methods study using a convergent parallel design [25]. The sample size for the primary endpoints was calculated to be a maximum of 160 participants, accounting for drop outs [24]. Study design and data analysis was carried out concurrently for the quantitative and qualitative elements. Comparison of the results allows for a deeper understanding of the participants’ experiences (Figure 1). Forty generally healthy families with two adults (18-60y) and between one and three children (28d-17y), living in or near Liverpool, United Kingdom, were recruited to collect samples of saliva and nasal lining fluid every two weeks at home for six months. To capture microbial transmission, participants were also asked to collect hand swabs at the same timepoints.

**Figure 1:**
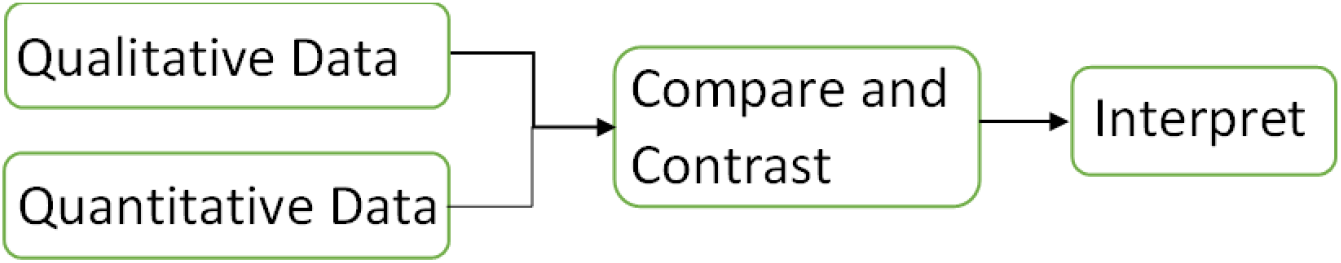
Convergent Parallel Design, adapted from [26]

Participants were asked to provide a saliva sample at least 30 minutes after last eating, drinking, or brushing their teeth. A sample of nasal fluid was collected by applying a soft synthetic absorptive matrix (SAM, Nasosorption^™^ FX-i, Mucosal Diagnostics) to the nasal lining for up to 2 minutes. Hand swabs were collected by swabbing the dominant hand with a wet swab. Samples were stored in domestic freezers until pick-up and transport to the laboratories at the Liverpool School of Tropical Medicine. Sample pick-ups were arranged half-way through and at the end of the study, unless more frequent pick-ups were requested by the participants.

The COVID-19 pandemic disrupted recruitment, with participants mainly recruited by word of mouth and through staff communications at local hospitals and universities, rather than through hospital outpatient lists as originally planned. Moreover, because of COVID-19 restriction measures, most appointments for screening, obtaining informed consent and training in sample collection took place virtually over Microsoft Teams. Communication with parents was maintained throughout the study via WhatsApp, including reminders of sampling dates, arranging times for sample pick-ups, and answering queries regarding the study.

### Assessing study experiences: quantitative

Families were asked to complete a questionnaire after every sampling timepoint to monitor each participant’s discomfort and pain individually using Likert scales. The Likert scales for discomfort were the same for adults and children, but the Likert scale for pain in children showed faces with different expressions to aid accuracy of results in young children. The practicality questionnaire also monitored the time taken to collect each sample type and the occurrence of nosebleeds when collecting nasal fluid (Supplementary1: Practicality questionnaire).

Questionnaire data was entered manually into Microsoft Excel and analysed using GraphPad Prism and R software.

### Assessing study experiences: qualitative

As this was a small pilot study, we used convenience sampling to attain data saturation, with a minimum sample size of 12 interviews with participants from different households [27]. All parents were given the opportunity to participate in an optional exit interview after study completion. Structured interviews (Supplementary2: Interview questions) were conducted virtually by a study researcher (EN) over Microsoft Teams, because of COVID-19 restriction measures. Interviews were recorded and transcribed automatically, after participants’ verbal consent, with transcription subsequently quality controlled by a second study researcher (ELG). Quality control entailed watching the recorded interviews frame by frame and correcting the transcribed text to reflect exactly what was said.

Corrected interview transcripts were used to identify key themes and to develop a coding framework with headings and subheadings (Supplementary2: coding framework) (ELG with HMN). Transcripts were uploaded into Nvivo 12 software and coded according to the framework agreed on (ELG). Parent identifiers were anonymised and the ones cited here are only known to the study team.

EN and ELG constituted the core study team, responsible for recruitment, training in sample collection, kit preparation, document management, liaising with participants and sample pick up, storage and laboratory analysis. Neither researcher had prior experience in qualitative research although EG had received training in qualitative methods. They received guidance from HMN who has extensive experience in qualitative research.

## Results

### Quantitative

36/41 families completed practicality questionnaires after sample collection, with data from a total of 141 participants (Table 1). Results from these questionnaires represented 92% of the total number of sampling events.

**Table 1:** Number of individuals and sampling events included in the quantitative analysis

| Group | Individuals included | Sampling events represented |
| --- | --- | --- |
| Adults | 72 | 777 |
| Children >5y | 33 | 352 |
| Children ≤5y | 36 | 382 |
| Total | 141 | 1540 |

#### Self-reported discomfort and pain

Overall, “no discomfort” was reported for 895 (58%, 95% CI [56%,61%]) sample collections: 576 (73%, 95% CI [71%,77%]) sample collections from adults, 171 (47%, 95% CI [43%,54%]) sample collections from children >5y and 148 (38%, 95% CI [34%,44%]) sample collections from children ≤5y. Moderate, severe, and extreme discomfort were reported during 128 (33%, 95% CI [29%,38%]) sample collections from children ≤5y compared to 26 (7%, 95% CI [5%,11%]) of sample collections from children >5y and 13 (2%, 95% CI [1%,3%]) of sample collections from adults (Figure 2).

**Figure 2:**
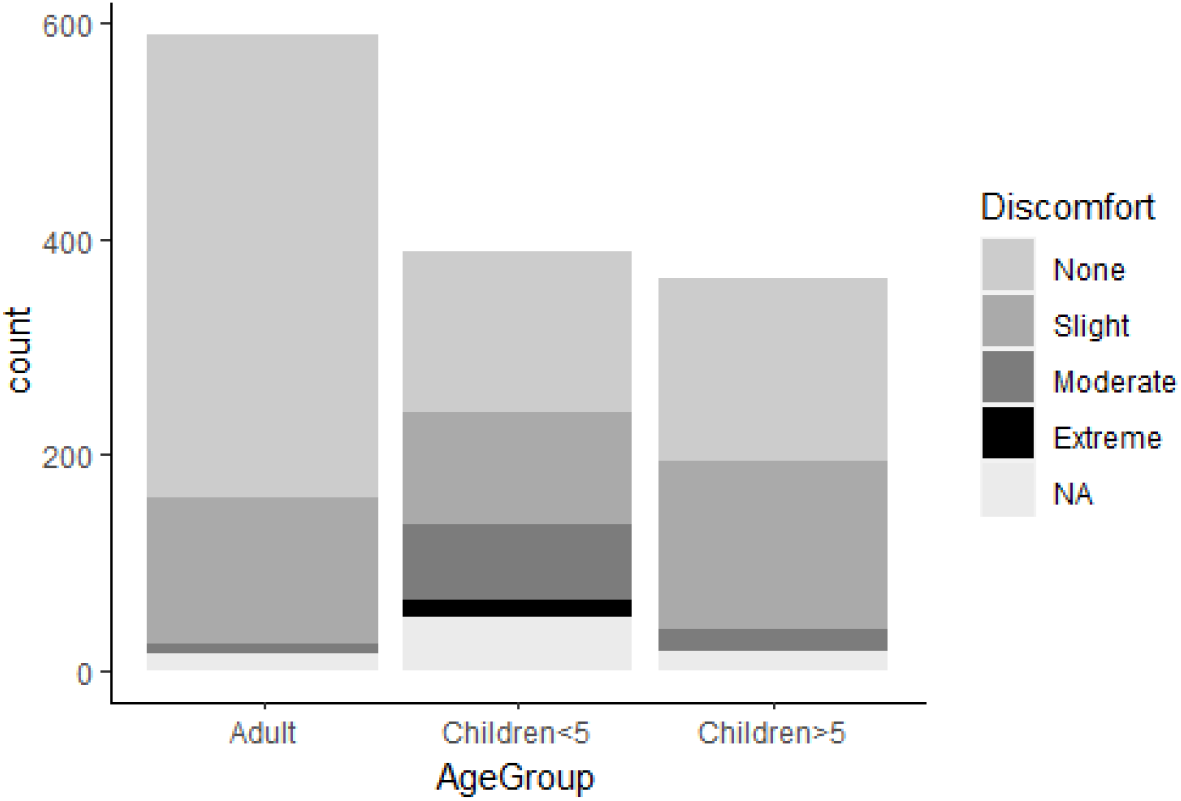
Discomfort scores in different participant age groups

Adults reported “no pain” in 740 (94%, 95% CI [93%,97%]) sample collections. No pain was reported in 256 (70%, 95% CI [68%,77%]) sample collections from children >5y and 187 (48%, 95% CI [44%,54%]) sample collections in children ≤5y. The highest pain scores (hurt even more, a whole lot or the worst) were reported in 53 (14%, 95% CI [11%,18%]) sampling events in children ≤5y compared to only one sampling event in children >5y (Figure 3).

**Figure 3:**
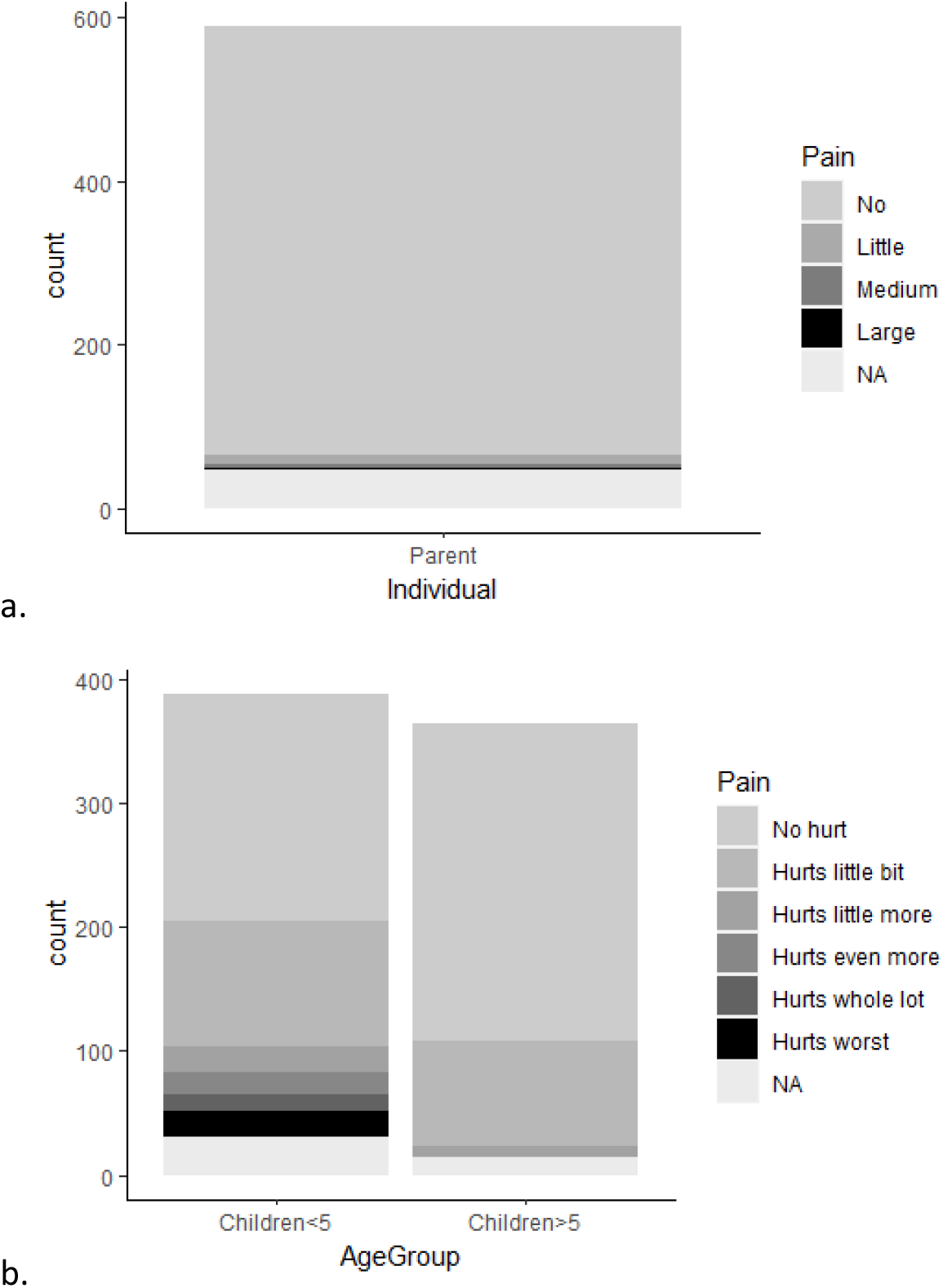
Pain scores in a) adults and b) children of different ages

Minor bleeding with nasal sampling was recorded in 59 instances (4%, 95% CI [3%,5%]), 33 times in adults (4%, 95% CI [3%,6%]) and 25 times in children (4%,95% CI [2%,5%]).

#### Time taken for sample collection

The mean time taken to collect samples differed by age group. Hand swabs and nasal sampling were completed faster for children ≤5y than for adults and children >5y. For saliva, it was children >5y that took the longest to sample. Average time taken to collect saliva decreased over the study period in all age groups: from 1.9min (SD=2.1) to 1.4min (SD=1.1) in adults, from 2.7min (SD=2.5) to 1.9min (SD=1.6) in children >5y and from 2.1min (SD=1.3) to 1.6min (SD=1.0) in children ≤5y. Likewise, average time taken to collect hand swabs also decreased from 0.9min (SD=1.0) to 0.6min (SD=0.4) in adults, from 0.9min (SD=1.0) to 0.6min (SD=0.5) in children >5y and from 0.6min (SD=0.3) to 0.4min (SD=0.2) in children ≤5y. Conversely, average time taken to collect nasal samples remained relatively constant around the requested 2min mark for adults and older children, and at an average of 1.3min for younger children (Figure 4).

**Figure 4:**
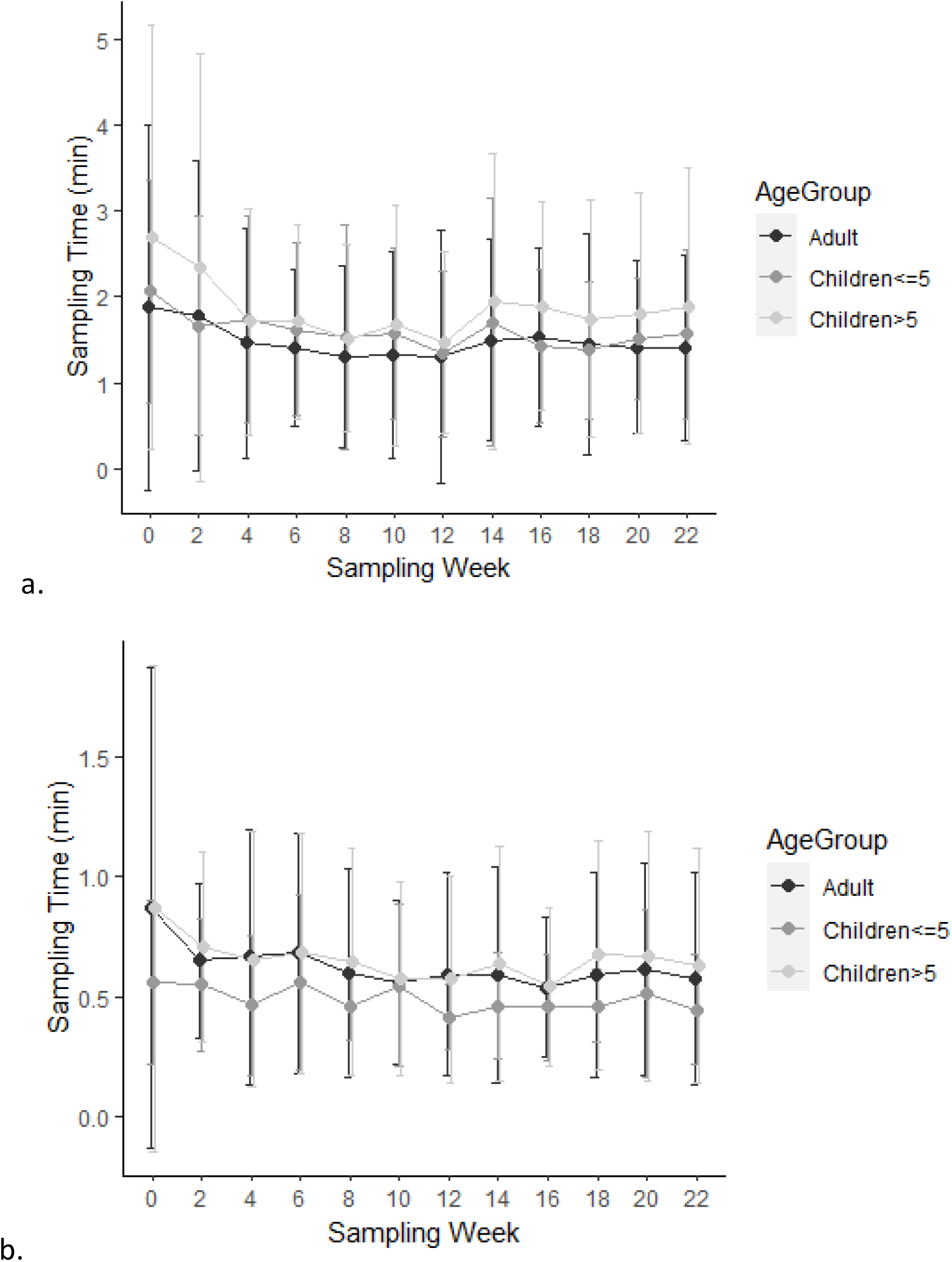
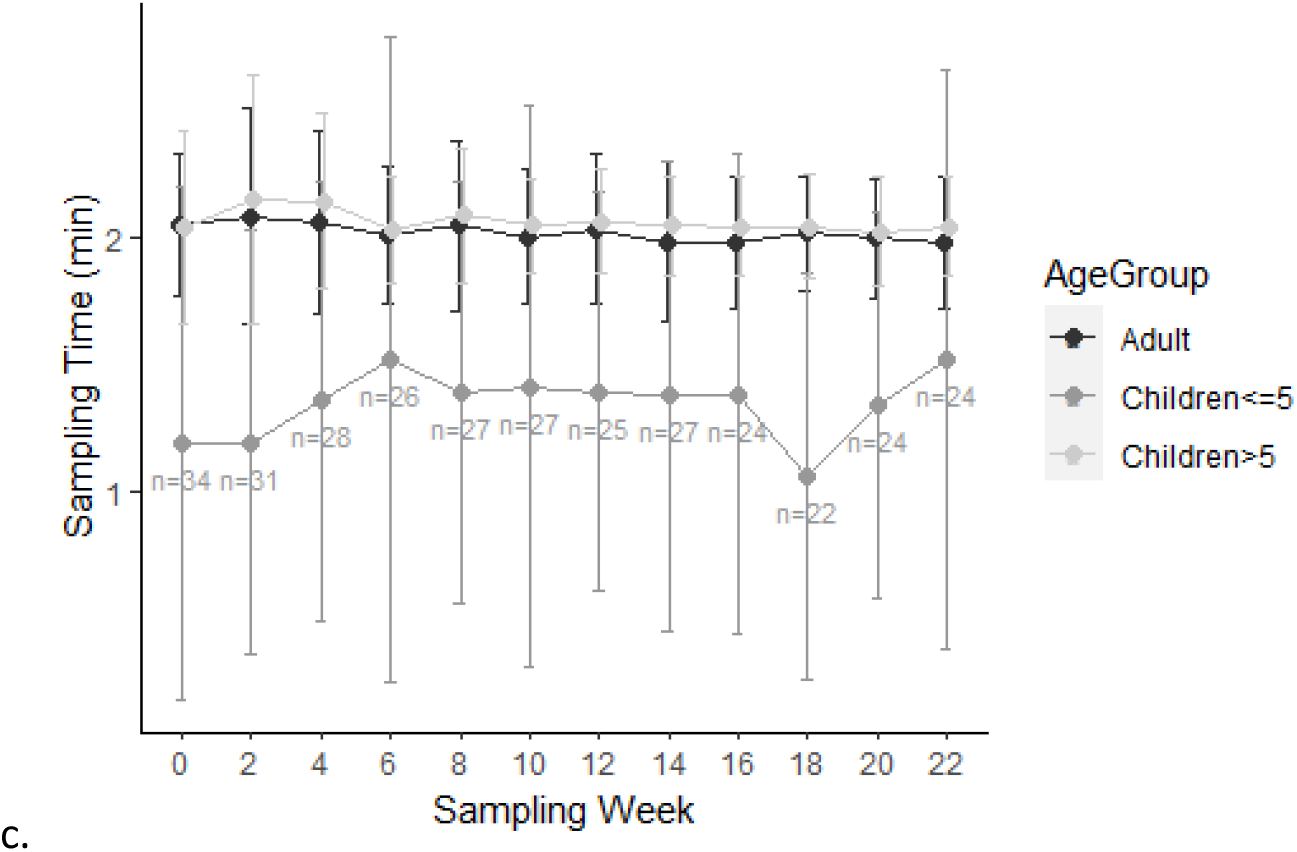
Timings of sample collection by age group and study week for a) Saliva, b) Hand swabs and c) Nasal sampling . Mean ± SD

### Qualitative

24 exit interviews with parents were conducted between March and September 2021, representing the experiences of 48 parents, 20 children >5y and 24 children ≤5y. There were 4 key themes i) Simplicity of procedure enhanced adherence ii) Preference for some sampling techniques was age dependent iii) Adequate time and fridge/freezer space is important for longitudinal sampling at home iv) Participant engagement with longitudinal sampling influenced by family’s motivation.

### Simplicity of procedure enhanced adherence

Parents found that the sampling method was simple to carry out.

*“Uhm ah, to be honest, we didn’t find anything particularly difficult. We thought it was very sort of straightforward.”* (Parent#49)

Initially, some parents found following the instructions and organising the sampling tubes complicated and overwhelming, however on repeated sampling they found it much easier. Parent employment in a clinical, research or scientific environment was advantageous for initial comprehension of the sampling protocols, but all families were able to collect samples correctly.

*“So yeah, the other thing is maybe first time again because I’m not in the field, it was very clear how to take samples but maybe I would have felt more confident in kind of demonstration when I was there like this is how children needs to chew […] Yeah, because I’m doing the covid stuff at home and I watched the video so that I think that same thing like you watch the video. You see it, you can do it, is yeah, it, it clears out any doubts.”* (Parent#21)

Parents cited both the training session at the start of the study and the laminated information sheet provided with the study pack as helpful for guiding sample collection. However, some would have preferred to have the training sessions in person rather than online, and to have the sample packs with them during the session to aid comprehension. In addition, some parents mentioned that having the information in video format would have been helpful.

*“I didn’t have this stuff in front of me when (the study team member) was explaining to me just due to the logistical issues, but I’m sure if we had in front of us it would have been easier but because of COVID and other things, but on the whole it was fine.”* (Parent#74)

### Preference for some sampling techniques was age dependent

Hand swabs were universally acknowledged as simple and comfortable to collect from all family members.

Saliva sample collection was straightforward for most participants, although a few young children did not co-operate, and a few teenagers and adults found the process “disgusting”. Conversely, other children really enjoyed collecting saliva samples because their parents made the procedure into a competitive game.

*“I didn’t particularly like doing the saliva samples, but just because if you think about it and you look at it, it was just a bit, just a bit gross.”* (Parent#71)

*“Oh yeah, the kids loved doing the saliva one, ‘cause they like, they liked to spit in the tub to get to the line. So that was their favourite one to do.”* (Parent#53)

Nasal sampling was generally easy for participants with the notable exception of the youngest children. Parents struggled to collect nasal samples from these children who sometimes became quite distressed, which in turn caused distress for the carers.

*“The nose, (child ≤5y) really didn’t like it. And we never, we never managed to do it for the full time requested just because he gets so upset, so we just do it for a little bit and stop. So, I don’t know how you get around that, or whether you just do it in older children who understand a bit more or something, but we had to like, restrain him and it, it wasn’t very nice.” (Parent#43)*

*“My eldest (≤5y) hated the nose. She absolutely hated it and she cried when we knew we were going to do it and she kept putting her hands over her face, she absolutely hated it.”* (Parent#64)

There were divergent views between children. Parent described some children as being “particularly sensitive”, therefore struggling with the procedure. Some children adjusted to the procedure over time whereas others started to refuse nasal sampling after a few weeks, or their parents gave up because of how challenging it was. Some parents did not think that the procedure was actually painful for their children, but rather that young children hate having anything in their noses and being restrained. They thought that sampling would be easier in babies and older children than in toddlers. Sampling during sleep was not an effective technique as the discomfort would often wake the child. In slightly older children, distraction using television and treats helped parents to encourage their children to participate in nasal sampling.

*“Yeah. Yeah, so if they (children ≤5y) had been much younger so if they had been tiny babies, it would have been fine to do it and I’d just hold them down and do it. And if they had been slightly older you know I’ll give you a chocolate button if you let me do this, but they just, but they just, there was nothing that could convince them to do it.”* (Parent#45)

*“I know there was a suggestion to do it when they’re sleeping, but for my baby, I walk in, she wakes up so I can’t. I can’t even approach her with that and the moment I just go near it, she smacks my hand so, yeah, she hates, she hates anything up the nose yeah so.”* (Parent#75)

Parents reported that previous sampling for SARS-Cov-2 using nasopharyngeal swabs predisposed their children to dislike nasosorption sampling.

*“I think also because of the pandemic and things, having, he’s (≤5y) had to have COVID swabs, you know, the cotton buds stuck up his nose by nurses? They’re not the most pleasant of experiences, so he’s developed a bit of a phobia about having things near his nose, so that won’t have helped.”* (Parent#15)

However, in older children and in adults, nasosorption sampling was compared favourably to swabbing for SARS-CoV-2.

*“Certainly, it’s a lot less upsetting than nasopharyngeal samples and our (child >5y) […] she really liked it as a method - she, she really engaged with it. She found it really, really easy.”* (Parent#01)

*“I’ll be honest, they (children >5y) didn’t love it, but they didn’t complain and they’re obviously doing the lateral flow tests at the minute because they’re in school. So, there’s no contrast is there? One’s deeply more unpleasant than the other.”* (Parent#69)

### Adequate time and fridge/freezer space is important for longitudinal sampling at home

Parents reported that the time required for sample collection appeared to get shorter over time. However, there were diverse parental attitudes on whether or not it was a reasonable amount of time to expect a family to take out of their schedules.

*“We got quicker at taking the samples as we got used to taking the samples. The first time we did it, it probably took about an hour ‘cause we weren’t familiar with what to do, but then you can probably do four people in less than half an hour if you coordinate it right now.” (Parent#15)*

*“But it was just, it became, it wasn’t just a kind of 10-minute job. It was like a as, a kind of, as a parent that was trying to do all, it took about, Maybe, sorry, about 30 minutes, 40 minutes in a day, which is quite a long time I suppose.”* (Parent#74)

Parents also reported that the timing of sample collection was challenging, because of the requirement to not have eaten, drunk or brushed their teeth in the half hour before collecting saliva. Although not a requirement, most families preferred to collect all samples at the same time to get it out of the way and so that both parents were on hand to help with younger children. This did increase the complexity of finding a time during the day for sampling.

*“Quite relieved it’s over to be honest, not having to think about it anymore, or. Especially now like traveling. I work at weekends and work has started up again. You know trying to think oh. And trying to get like the whole family to do it together like has everyone not eaten for half an hour you know or cleaned your teeth or with a child who constantly snacks or whatever, so it just took brainpower that I didn’t particularly want to do.”* (Parent#43)

Storage of study materials at home was an issue for some families with small fridges/freezers and especially during the holidays, as the sample collection tubes occupy a significant amount of space. However, most parents commented that while they agreed this could be a challenge, they themselves were not greatly inconvenienced. Parents were happy for the sample pick up service via taxi that was provided on demand, with a few suggestions of modifications such as staggered delivery of consumables throughout the study and more frequent, pre-scheduled pick-ups.

*“So we found that quite an imposition. Um, you know we had a, we had, I was gonna say arguments, discussions about whether we should turn on our our our Christmas freezer or not to keep these samples going or whatever. Otherwise, we just buy less frozen food. So yeah, […] I think when you’re getting to kind of two months, it’s it’s quite a lot of stuff in your freezer.”* (Parent#19)

*“We’re quite lucky, we’ve got, we’ve got freezer space that’s not a problem, but I can imagine if you’re short on freezer space then that might be a problem because you know each each of the saliva bottles was, you know it was they were. They were quite big.”* (Parent#71)

### Participant engagement with longitudinal sampling influenced by family’s motivation

Although the majority of parents said they could have easily continued the study for longer, some struggled to maintain motivation over the six-month period. Turning sample collection into a game or competition, and rewarding participation were techniques that worked with many children. Another key element for perseverance with the study was interest in and perceived importance of the research.

*“My boys definitely didn’t love doing it, but I’m hoping that there’s some value that comes out of it in terms of, you know, understanding how transmission so for me it was feeling like it was possibly quite useful in the long term.”* (Parent#69)

*“uhm the kids actually really enjoyed doing it. They were like “Yay!” every, every two weeks they like really enjoyed doing it. It was really, it was only my husband who was like “ah” and I was like “just do it” after like you know uhm a couple of weeks. He was the one who was like “uh can’t be bothered!”“* (Parent#53)

The use of WhatsApp messaging was universally appreciated as a simple and convenient way to maintain contact between the families and the study team.

*“I find it like if I had to ring somewhere, I’d find that more inconvenient because it’s more effort when you’ve got kids and when you’re out and about and you don’t stop, so that would have been inconvenient, but just a quick message is fine then you can pick it up when you next see your phone and uhm so that was, that was really easy.”* (Parent#53)

*“I didn’t really need to contact anybody, but I liked the fact that I got reminders to remind me about the samples ‘cause you do sometimes forget. And I knew that there was somebody if I needed them.”* (Parent#67)

## Discussion

We used mixed methods to gain an in depth understanding of the experiences and perceptions of families taking part in a longitudinal community-based respiratory sampling study. Parents reported that sampling was a relatively straightforward procedure that they quickly got used to carrying out but this was dependent on clear written and verbal instructions, as well as on the parent’s professional background. Sampling was quick in all age groups and the time decreased with experience for saliva and hand swabs. The recommended 2 minutes of nasal sampling was achieved in adults and older children, but the average time taken for nasal sampling for children ≤5y remained lower. Although the average time taken for nasal sampling is significantly different between samples that were positive or negative for the presence of *Streptococcus pneumoniae*, positive samples was associated with a shorter sampling time (data not shown). We hypothesise this is because children, especially ≤5y, have higher carriage rates. We therefore believe that short sampling times in young children still yield good results. Despite the speed and simplicity of the procedure, logistical challenges remained - families found it challenging to fit it around their busy schedules because of several pre-requisites, and there is also a requirement for considerable fridge/freezer space.

The results of the questionnaire showed that most participants of all ages found the minimally invasive sampling method to be comfortable and painless, though a significant proportion of children ≤5y did find the procedure uncomfortable and painful. Interviews further clarified that this discomfort and pain was mainly caused by the nasal sampling. Although nasal sampling using a synthetic absorptive matrix compared favourably with nasopharyngeal swabs in adults and older children, they were more challenging for the youngest children who disliked being restrained and having anything in their noses. Although synthetic absorptive matrices have been used successfully in studies of hospitalised children under anaesthesia [28, 29] and in home sampling in adults [23], to our knowledge, this study is the first time they have been used by parents to collect samples in children in a community setting.

Despite the challenges above, participant compliance with the sampling schedule was very high in both adults and children [24]. Using the WhatsApp social media platform was reported as being very successful for maintaining good communication with participants, and study team engagement with them was helpful for boosting motivation.

The key strength of this analysis is the quantity of data collected. Most participating families completed both regular practicality questionnaires and the exit interview. The key messages were consistent across the interviews, suggesting that we achieved data saturation while conducting the qualitative interviews. Furthermore, the data collected reflects the experience of children across a wide range of ages, allowing for distinction between age groups.

There are limitations to our findings. Firstly, interviews were conducted with parents, usually only the most involved parent, and therefore probably did not reflect the full opinions of all family members. Likewise, getting an accurate assessment of the levels of pain and discomfort in young children, particularly infants, is challenging, and the responses will reflect parents’ interpretation of distress levels which can be very subjective [10, 11]. This was evident in our study where some, but not all, parents with younger children stopped collecting nasal lining fluid as they felt that their children were too distressed [24]. Using a visual scoring system for discomfort with faces showing different expressions may have helped standardise reporting for toddlers and pre-schoolers who cannot yet read. EN and ELG were the primary researchers conducting the FAMILY Micro study and neither have a background in qualitative research. The expertise of HMN was solicitated to minimise any resulting bias or tendency to approach data analysis quantitatively, but some may remain. Using semi-structured interviews with inductive probing rather than a fully structured format may have increased the depth of our results. There are disadvantages to virtual interviewing, such as limited visibility of body language cues or increased distractions from the home environment. However, research suggests that researcher-interviewee rapport and data richness are similar between virtual and face-to-face interviews [30].

Due to changes in recruitment strategy due to COVID-19 restriction measures, the study cohort included a high proportion of medical practitioners and researchers. Furthermore, this cohort had higher income and education levels than the national average despite the Liverpool being one of the most deprived local authorities in the UK [31]. They would therefore have been more likely to understand, engage with and comply with study activities than the general population. It is also possible that reduced activities outside of the home because of COVID-19 restrictions (such as home-schooling, work from home, no holidays, no extracurricular activities, no visiting… etc) increased the likelihood of participants persevering with study activities.

## Conclusion

With a few adjustments such as video tutorials and staggered delivery of consumables, we believe that, overall, our home sampling method is feasible for long-term community surveillance in families. Although the use of nasosorption devices rather than nasopharyngeal swabs was well-received by adults and older children, it did not resolve the issue of discomfort during nasal sampling in children ≤5y. More research is needed in this group as they are critically important for respiratory disease burden and pathogen transmission.

## Data Availability

All data produced in the present study are available upon reasonable request to the authors

## Acknowledgements

We would like to thank Kelly Davies and the LSTM research governance team for their assistance in setting up the FAMILY Micro study. We thank Alder Hey children’s hospital for their collaboration as a study recruitment site. We are also grateful to the many volunteers who helped label sample collection tubes and storage containers. Most of all though, we would like to thank the study participants who gave so generously of their time and commitment during an extremely challenging year. The study was supported by the Director’s Catalyst Fund 2020 awarded to Dr Elissavet Nikolaou and the UKRI Strength in Places Fund awarded to Prof Ferreira.

## Notes

### Competing Interest Statement

The authors have declared no competing interest.

### Funding Statement

The study was supported by the Directors Catalyst Fund 2020 awarded to Dr Elissavet Nikolaou and the UKRI Strength in Places Fund awarded to Prof Ferreira

### Author Declarations

The North West - Greater Manchester West Research Ethics Committee gave ethical approval for this work (20/NW/0304)

## References

1. Strong, K.L., et al., Patterns and trends in causes of child and adolescent mortality 2000– 2016: setting the scene for child health redesign. BMJ Global Health, 2021. 6(3): p. e004760.

2. Liu, L., et al., Global, regional, and national causes of child mortality in 2000–13, with projections to inform post-2015 priorities: an updated systematic analysis. The Lancet, 2015. 385(9966): p. 430–440.

3. Galanti, M., et al., Rates of asymptomatic respiratory virus infection across age groups. Epidemiol Infect, 2019. 147: p. e176.

4. Weiser, J.N., D.M. Ferreira, and J.C. Paton, Streptococcus pneumoniae: transmission, colonization and invasion. Nat Rev Microbiol, 2018. 16(6): p. 355–367.

5. Sundell, N., et al., PCR Detection of Respiratory Pathogens in Asymptomatic and Symptomatic Adults. J Clin Microbiol, 2019. 57(1).

6. DeMuri, G.P., et al., Dynamics of Bacterial Colonization With Streptococcus pneumoniae, Haemophilus influenzae, and Moraxella catarrhalis During Symptomatic and Asymptomatic Viral Upper Respiratory Tract Infection. Clin Infect Dis, 2018. 66(7): p. 1045–1053.

7. Jansen, R.R., et al., Frequent detection of respiratory viruses without symptoms: toward defining clinically relevant cutoff values. J Clin Microbiol, 2011. 49(7): p. 2631–6.

8. Self, W.H., et al., Respiratory Viral Detection in Children and Adults: Comparing Asymptomatic Controls and Patients With Community-Acquired Pneumonia. J Infect Dis, 2016. 213(4): p. 584–91.

9. Singleton, R.J., et al., Viral respiratory infections in hospitalized and community control children in Alaska. J Med Virol, 2010. 82(7): p. 1282–90.

10. Kawade, A., et al., Assessment of perceived distress due to nasopharyngeal swab collection in healthy Indian infants participating in a clinical trial. Paediatr Neonatal Pain, 2021. 3(4): p. 170–175.

11. Westra, A.E., et al., Perceived discomfort levels in healthy children participating in vaccine research. J Empir Res Hum Res Ethics, 2013. 8(3): p. 66–72.

12. Gagnon, F., et al., Nasopharyngeal swabs vs. saliva sampling for SARS-CoV-2 detection: A cross-sectional survey of acceptability for caregivers and children after experiencing both methods. PLoS One, 2022. 17(7): p. e0270929.

13. Lambert, S.B., K.M. Allen, and T.M. Nolan, Parent-collected respiratory specimens--a novel method for respiratory virus and vaccine efficacy research. Vaccine, 2008. 26(15): p. 1826–31.

14. Esposito, S., et al., Collection by trained pediatricians or parents of mid-turbinate nasal flocked swabs for the detection of influenza viruses in childhood. Virol J, 2010. 7: p. 85.

15. Akmatov, M.K. and F. Pessler, Self-collected nasal swabs to detect infection and colonization: a useful tool for population-based epidemiological studies? Int J Infect Dis, 2011. 15(9): p. e589–93.

16. Akmatov, M.K., et al., Equivalence of self- and staff-collected nasal swabs for the detection of viral respiratory pathogens. PLoS One, 2012. 7(11): p. e48508.

17. Coughtrie, A.L., et al., Evaluation of swabbing methods for estimating the prevalence of bacterial carriage in the upper respiratory tract: a cross sectional study. BMJ Open, 2014. 4(10): p. e005341.

18. Nguyen, T.T., et al., Accuracy and Acceptance of a Self-Collection Model for Respiratory Tract Infection Diagnostics: A Concise Clinical Literature Review. J Emerg Nurs, 2021. 47(5): p. 798–806.

19. Miellet, W.R., et al., Detection of Neisseria meningitidis in saliva and oropharyngeal samples from college students. Sci Rep, 2021. 11(1): p. 23138.

20. Wyllie, A.L., et al., Streptococcus pneumoniae in saliva of Dutch primary school children. PLoS One, 2014. 9(7): p. e102045.

21. Byrne, R.L., et al., Saliva Alternative to Upper Respiratory Swabs for SARS-CoV-2 Diagnosis. Emerg Infect Dis, 2020. 26(11): p. 2770–2771.

22. Yasuda, I., et al., Respiratory virus detection in the upper respiratory tract of asymptomatic, community-dwelling older people. BMC Infect Dis, 2022. 22(1): p. 411.

23. Nikolaou, E., et al., Experimental Human Challenge Defines Distinct Pneumococcal Kinetic Profiles and Mucosal Responses between Colonized and Non-Colonized Adults. mBio, 2021. 12(1).

24. Nikolaou, E., et al., Family Research of Microbes Linked to Respiratory Infections (FAMILY Micro) observational study: Assessing the use of minimally invasive self-sampling methodologies at home for long-term monitoring of the oral, nasal and hand microbiota of adults and children within UK families. medRxiv, 2023: p. 2023.04.19.23288393.

25. Creswell, J. and J. Creswell, Research Design: Qualitative, Quantitative and Mixed Methods Approaches. 5th ed. 2017: SAGE publications, Inc.

26. Creswell, J. and V. Plano Clark, Designing and Conducting Mixed Methods Research. 2nd Edition ed. 2011: Sage Publications.

27. Guest, G., A. Bunce, and L. Johnson, How Many Interviews Are Enough?:An Experiment with Data Saturation and Variability. Field Methods, 2006. 18(1): p. 59–82.

28. Thwaites, R.S., et al., Nasosorption as a Minimally Invasive Sampling Procedure: Mucosal Viral Load and Inflammation in Primary RSV Bronchiolitis. J Infect Dis, 2017. 215(8): p. 1240–1244.

29. Nikolaou, E., et al., Minimally invasive nasal sampling in children offers accurate pneumococcal colonization detection. Pediatr Infect Dis J, 2019. 38(11): p. 1147–1149.

30. Keen, S., M. Lomeli-Rodriguez, and H. Joffe, From Challenge to Opportunity: Virtual Qualitative Research During COVID-19 and Beyond. Int J Qual Methods, 2022. 21: p. 16094069221105075.

31. Council, L.C., The Index of Multiple Deprivation 2019: A Liverpool analysis. 2020.

