## Supplemental Material 1 for "Participant perceptions and experiences of a novel community-based pilot respiratory longitudinal sampling method in Liverpool, UK"

### Home Sampling Practicality Questionnaire

**TO BE COMPLETED AT EVERY STUDY DATE AFTER SAMPLE COLLECTION**

Instructions: Please read the questions carefully and record your answers below. To be completed by all family members (parents can record for children).

1. Did you feel any discomfort (uncomfortable) taking the sample? Indicate your level of discomfort by ticking one box only ☑

***PARENT A***

| I felt: | Strongly disagree | Disagree | Neither agree or disagree | Agree | Strongly Agree |
|---|---|---|---|---|---|
| **No** discomfort | ☐ | ☐ | ☐ | ☐ | ☐ |
| **Slight** discomfort | ☐ | ☐ | ☐ | ☐ | ☐ |
| **Moderate** discomfort | ☐ | ☐ | ☐ | ☐ | ☐ |
| **Severe** discomfort | ☐ | ☐ | ☐ | ☐ | ☐ |
| **Extreme** discomfort | ☐ | ☐ | ☐ | ☐ | ☐ |

***PARENT B***

| I felt: | Strongly disagree | Disagree | Neither agree or disagree | Agree | Strongly Agree |
|---|---|---|---|---|---|
| **No** discomfort | ☐ | ☐ | ☐ | ☐ | ☐ |
| **Slight** discomfort | ☐ | ☐ | ☐ | ☐ | ☐ |
| **Moderate** discomfort | ☐ | ☐ | ☐ | ☐ | ☐ |
| **Severe** discomfort | ☐ | ☐ | ☐ | ☐ | ☐ |
| **Extreme** discomfort | ☐ | ☐ | ☐ | ☐ | ☐ |

***CHILD A***

| I felt: | Strongly disagree | Disagree | Neither agree or disagree | Agree | Strongly Agree |
|---|---|---|---|---|---|
| **No** discomfort | ☐ | ☐ | ☐ | ☐ | ☐ |
| **Slight** discomfort | ☐ | ☐ | ☐ | ☐ | ☐ |
| **Moderate** discomfort | ☐ | ☐ | ☐ | ☐ | ☐ |
| **Severe** discomfort | ☐ | ☐ | ☐ | ☐ | ☐ |
| **Extreme** discomfort | ☐ | ☐ | ☐ | ☐ | ☐ |

***CHILD B***

| I felt: | Strongly disagree | Disagree | Neither agree or disagree | Agree | Strongly Agree |
|---|---|---|---|---|---|
| **No** discomfort | ☐ | ☐ | ☐ | ☐ | ☐ |
| **Slight** discomfort | ☐ | ☐ | ☐ | ☐ | ☐ |
| **Moderate** discomfort | ☐ | ☐ | ☐ | ☐ | ☐ |
| **Severe** discomfort | ☐ | ☐ | ☐ | ☐ | ☐ |
| **Extreme** discomfort | ☐ | ☐ | ☐ | ☐ | ☐ |

***CHILD C***

| I felt: | Strongly disagree | Disagree | Neither agree or disagree | Agree | Strongly Agree |
|---|---|---|---|---|---|
| **No** discomfort | ☐ | ☐ | ☐ | ☐ | ☐ |
| **Slight** discomfort | ☐ | ☐ | ☐ | ☐ | ☐ |
| **Moderate** discomfort | ☐ | ☐ | ☐ | ☐ | ☐ |
| **Severe** discomfort | ☐ | ☐ | ☐ | ☐ | ☐ |
| **Extreme** discomfort | ☐ | ☐ | ☐ | ☐ | ☐ |

2. Did you feel any pain taking the sample? Indicate your level of pain by ticking one box only ☑

***PARENT A***

| I felt: | Strongly disagree | Disagree | Neither agree or disagree | Agree | Strongly Agree |
|---|---|---|---|---|---|
| **No** pain | ☐ | ☐ | ☐ | ☐ | ☐ |
| **Little** pain | ☐ | ☐ | ☐ | ☐ | ☐ |
| **Medium** pain | ☐ | ☐ | ☐ | ☐ | ☐ |
| **Large** pain | ☐ | ☐ | ☐ | ☐ | ☐ |
| **Worst possible** pain | ☐ | ☐ | ☐ | ☐ | ☐ |

***PARENT B***

| I felt: | Strongly disagree | Disagree | Neither agree or disagree | Agree | Strongly Agree |
|---|---|---|---|---|---|
| **No** pain | ☐ | ☐ | ☐ | ☐ | ☐ |
| **Little** pain | ☐ | ☐ | ☐ | ☐ | ☐ |
| **Medium** pain | ☐ | ☐ | ☐ | ☐ | ☐ |
| **Large** pain | ☐ | ☐ | ☐ | ☐ | ☐ |
| **Worst possible** pain | ☐ | ☐ | ☐ | ☐ | ☐ |

***CHILD A***

☐ ☐ ☐ ☐ ☐ ☐

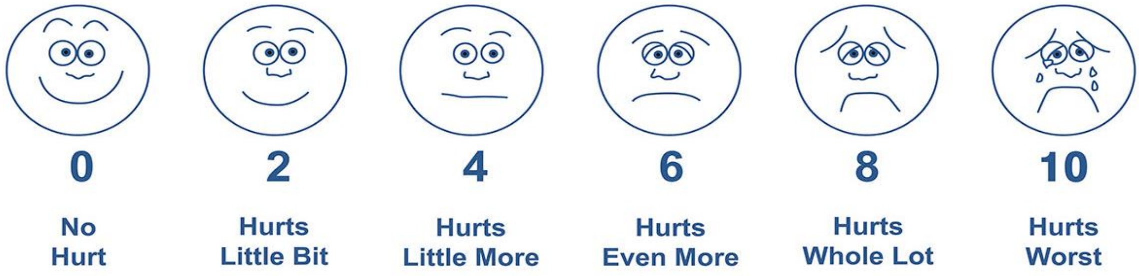

***CHILD B***

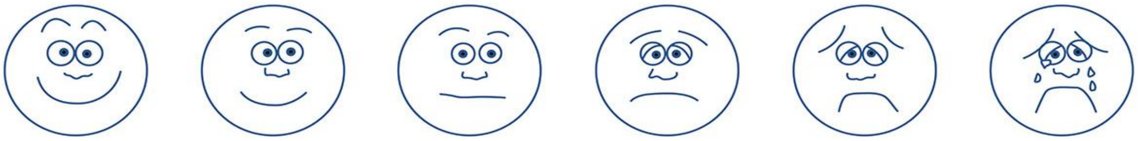

| 0 | 2 | 4 | 6 | 8 | 10 |
|---|---|---|---|---|---|
| No Hurt | Hurts Little Bit | Hurts Little More | Hurts Even More | Hurts Whole Lot | Hurts Worst |
| ☐ | ☐ | ☐ | ☐ | ☐ | ☐ |

***CHILD C***

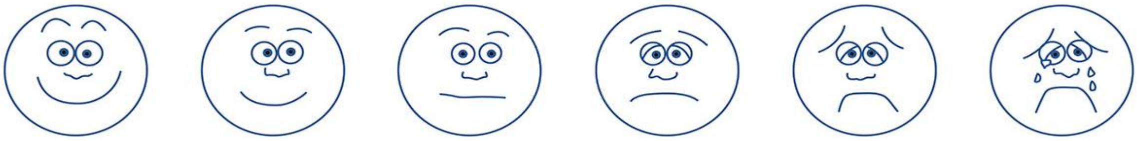

| 0 | 2 | 4 | 6 | 8 | 10 |
|---|---|---|---|---|---|
| No Hurt | Hurts Little Bit | Hurts Little More | Hurts Even More | Hurts Whole Lot | Hurts Worst |
| ☐ | ☐ | ☐ | ☐ | ☐ | ☐ |

3. Was there any bleeding while taking the nasal fluid (strip) sample (tick one box only ☑)?

| ***PARENT A*** | ***PARENT B*** | ***CHILD A*** | ***CHILD B*** | ***CHILD C*** |
|---|---|---|---|---|
| ☐Yes ☐No | ☐Yes ☐No | ☐Yes ☐No | ☐Yes ☐No | ☐Yes ☐No |

4. *For parents:* How long did it take you to collect the samples (record time e.g. 30sec, 1min, 2min)?

***PARENT A***

Time to collect SALIVA: ……………………………………………………………………………………………………

Time to collect NASAL FLUID (paper strip): ……………………………………………………………………..

Time to collect HAND SWAB: ………………………………………………………………………………………….

NOTES:

***PARENT B***

Time to collect SALIVA: ……………………………………………………………………………………………………

Time to collect NASAL FLUID (paper strip): ……………………………………………………………………..

Time to collect HAND SWAB: ………………………………………………………………………………………….

NOTES:

5. *For children:* How long did it take you to collect the samples from children (record time e.g. 30sec, 1min, 2min)? Please also indicate which device did you use for saliva (tube or swab) in each child.

***CHILD A***

Time to collect SALIVA: …………………………… Saliva device: ☐ Saliva tube ☐ Swab

Time to collect NASAL FLUID (paper strip): ………………………………………………………………

Time to collect HAND SWAB: ……………………………………………………………………………………

NOTES:

***CHILD B***

Time to collect SALIVA: ……………………………. Saliva device: ☐ Saliva tube ☐ Swab

Time to collect NASAL FLUID (paper strip): ……………………………………………………………….

Time to collect HAND SWAB: …………………………………………………………………………………….

NOTES:

***CHILD C***

Time to collect SALIVA: ……………………………. Saliva device: ☐ Saliva tube ☐ Swab

Time to collect NASAL FLUID (paper strip): ……………………………………………………………….

Time to collect HAND SWAB: …………………………………………………………………………………….

NOTES:
