## Supplemental Material 2 for "Participant perceptions and experiences of a novel community-based pilot respiratory longitudinal sampling method in Liverpool, UK"

### Semi-structured interview guide

**TO BE CONDUCTED AT THE END OF THE STUDY**

- Introduce yourself
- Go through the information leaflet and consent form
- Go over areas to cover
- Explain that you will note down anything that you want to come back to
- Reiterate that there is no right or wrong answer
- That the discussion will be recorded to facilitate recollection
- That all the information they give will be kept confidential and will only be shared with those involved in the study
- That the data collected will be anonymised so they will not be identified

Introduction: I have come to talk to you about your experience of home sampling to get a better understanding of how easy or difficult this is for parents and their children. It will take 10-15 minutes.

1. How was the home sampling procedure explained to you?
   - Was it useful?
   - How could it be improved?
2. How did you find the home sample information?
   - Was it easy to read?
   - Was it easy to understand?
3. How did you find the staff involved in this home sampling process?
   - Were they approachable?
   - Were they contactable?
4. How did you find the procedure?
   - What were the most challenging aspects for you?
   - What were the most challenging aspects for your children?
5. What could have been improved?
6. Please comment on your overall experience with home sample collection
7. As we aim to conduct a larger study in the future, we would like your opinion/input on the study design.
   - Do you like the name of the study?
   - Are the questionnaires/monthly diary instructions easy to follow?
   - Do you have any suggestions for improving the design of the study?
