## Supplemental Material 3 for "Participant perceptions and experiences of a novel community-based pilot respiratory longitudinal sampling method in Liverpool, UK"

**Coding framework for FAMILY Micro Study interviews**

- Favourable opinions
- Discomfort
    - Nasosorption samples
    - Saliva samples
    - Effect of Age
    - Impact of COVID swabs on this experience
        - Positive
        - Negative
- Disgust
    - Saliva samples
    - Effect of Age
- Inconvenience
    - Time taken
    - Disruption to family life
    - Freezer space
    - Length of study & maintaining motivation: effect of Covid
- Lack of clarity
    - Initial information: effect of Covid
    - Effect of occupation
    - Questionnaire wording
